# The Relevance of Lactate Levels in Acute Seizure

**DOI:** 10.1101/2022.02.04.22270161

**Authors:** Jessica Sop, Jessica Rogers, Nnennaya Opara, Alfred Tager, Scott Dean, Mark L Gustafson

**Affiliations:** Emergency Medicine, Charleston Area Medical Center, Charleston, USA; CAMC Education and Research Institute, Charleston Area Medical Center, Charleston, USA; Internal Medicine, Charleston Area Medical Center, Charleston, USA

**Keywords:** Lactate, Lactic Acid, Seizure, Epilepsy, Length of Stay

## Abstract

**Introduction:** Seizures can result in profound elevations of serum lactate. A paucity of investigation into whether lactate levels in these patients is associated with increased mortality. We sought to evaluate the significance in patients presenting with a seizure and elevated lactate.

**Methods:** This is a retrospective study involving patients presenting to the Emergency Department (ED) with a diagnosis of seizure from September 1st, 2016 until March 31st, 2018. Those with normal serum lactate (< 2mmol/L) were compared with patients that had an elevated serum lactate (≥ 2mmol/L).

**Results:** Patients with normal serum lactate levels had a significantly longer length of stay in the ED (60 minutes p = 0.04) when compared to those with elevated serum lactate levels. Additionally, a higher percentage of patients with elevated serum lactate received intravenous fluids in the ED (54.69% vs. 35.4%, P=0.01). There was no significant difference in other measured variables.

**Conclusion:** This study illustrates that serum lactate levels do not directly correlate with indicators of disease severity or outcomes when elevated secondary to seizure. However, patients with normal serum lactate levels had a longer length of stay in the ED.

## Introduction

A seizure is a sudden surge of electrical activity in the brain ^1^. The electrical activity that occurs in the human brain during seizures are caused by complex chemical alteration in the nerve cells. Seizures in of themselves are not a disease, rather, they are a symptom of a disorder that is affecting the brain. While some seizures occur unnoticed, others are totally disabling. Acute seizure accounts for about 1 million ER visits annually, which is approximately 1% of total ER visits ^2^. A systematic review of US healthcare costs for patients with seizure disorder revealed that epilepsy-specific costs ranged from $1,022 to $19,749 per person annually ^3^.

Serum lactate has been used as a predictor for risk of morbidity and mortality in many disease states. It has been widely used as an indicator of disease severity and mortality in sepsis and trauma ^4, 5^. Lactate has also been known to dramatically increase in the presence of acute seizure ^6^. Pre-existing medical conditions, seizure type, duration and intensity influence the resultant physiologic effects’ muscle contractions, increased oxygen demand and impaired breathing instigate temporary tissue hypoxia and hypo-perfusion during a tonic-clonic seizure ^7^.

Lactate is generated during this transient state of hypoxia as a product of anaerobic metabolism of glucose ^8^. Studies as early as 1977 have illustrated that lactic acidosis associated with seizure is self-limited and resolves even without intervention with a half-life of approximately 50% at 1 hour. Hence, when there is no other underlying cause for lactic acidosis, seizures can be considered a self-resolving lactic acidosis state ^9, 10^. It is a consequence of cessation of source production when the seizure stops ^11^. Clinical symptoms such as bite on tongue, drowsiness, incontinence all may hint at epileptic seizure and can also occur in non-epileptic seizures. This makes some diagnostic examinations such as computer tomography of head (Head CT), or Electro encephalography (EEG) unreliable in the diagnosis of seizures. Thus, making the laboratory markers such as creatinine kinase, lactic acid and prolactin very useful as they are all increased after an epileptic tonic-clonic seizure episode ^12^. Additionally, blood pH generally normalizes within 1 hour, and so, despite significant lactic acidosis, severe metabolic acidemia is not expected following the postictal recovery period subsequent to a generalized tonic-clonic seizures ^13, 14^. Therefore, persistent lactic acidosis beyond 1-2 hours following a seizure warrants further investigation ^9, 10, 15^.

## Materials and Methods

An observational, retrospective study was approved, and ethical approval (IRB: 18-444) was issued by the Charleston Area Medical Centre Institutional Review Board. Data were collected from patients who presented to the emergency department with the primary diagnosis of seizure. The data collection period was from September 12, 2017 through March 31, 2018.

Inclusion criteria for this study were patients who had the primary diagnosis of seizure and were ≥ 18 years of age. Patients were excluded if they were under 18 years of age or sustained trauma. Additionally, pregnant patients, end-stage renal disease patients, and patients taking metformin were eliminated. Furthermore, patients with an elevated ethanol level were excluded because of its ability to skew relative outcomes with regard to lactate elevation ^16^.

A serum lactate level of < 2 mmol/L was set as normal and anything above it was considered an elevated lab value ^17^. The measured variables, including sex, median age, antibiotic use, urine drug screen performed, Use of anti-epileptic medication, fluid administration, number of comorbidities and rate of admission, were compared between the two study groups as exhibited in table-1and table-2.

**Table-1:** Descriptive Information regarding study groups

| Variables | LA < 2 mmol/L<br>(N=115) | LA >2 mmol/L<br>(N=64) |
| --- | --- | --- |
| Gender (male & female) | 50 (43.5%) | 29 (45.3%) |
| Age – Median (Range) | 40 (18-80) | 38 (18-82) |
| Antibiotic Use | 4 (3.9%) | 2 (3.2%) |
| Urine Drug Screen | 20 (19.8%) | 15 (25.9%) |
| Anti- Seizure Meds Use | 80 (72.1%) | 36 (58.1%) |
| Total amount of Fluids<br>Given | 40L (35.4%) | 35L (54.7%) |
| **# of Comorbidities** |  |  |
| 0 | 30 (26.1%) | 27 (42.2%) |
| 1 | 49 (42.6%) | 15 (23.4%) |
| 2 | 22 (19.1%) | 14 (21.9%) |
| 3 | 10 (8.7%) | 3 (4.7%) |
| 4 | 2 (1.7%) | 4 (6.3%) |
| 5 | 2 (1.7%) | 1 (1.6%) |
| Admission rate | 9 (7.8%) | 3 (4.7%) |

LA: lactic acid

**Table-2:** percentage of other Comorbidities in the Group.

| Comorbidities | LA < 2 mmol/L<br>(N=11.5) | LA>2 mmol/L<br>(N=64) | P-Value |
| --- | --- | --- | --- |
| Hypertension | 19.1% | 12.5% | 0.255 |
| Diabetes Mellitus | 4.4% | 7.8% | 0.333 |
| Congestive Heart failure | 0.9% | 0% | 1 |
| Chronic Obstructive Pulmonary Disease. | 2.6% | 4.7% | 0.668 |
| Active Malignancy | 6.1% | 6.3% | 1 |
| Coronary Artery Disease | 1.7% | 4.7% | 0.3505 |
| Deep Venous Thrombosis | 1.7% | 4.7% | 0.3505 |
| Obesity | 15.7% | 9.4% | 0.2375 |
| Seizure Disorder | 58.3% | 48.4% | 0.2057 |
| Drug Addiction | 3.5% | 6.3% | 0.459 |
| Alcoholic | 7.8% | 6.3% | 0.7735 |
| Other | 0.9% | 3.1% | 0.291 |

LA: lactic acid

When IV fluids were administered, isotonic fluid was used. As noted, some patients received antibiotics in the ED if they were suspected to have an underlying infection causing their seizure. A urine drug screen was performed on patients who were thought to have used an illicit substance based on history and physical exam. Because we found that the majority of patients had a history of at least one medical condition, number of comorbidities was a parameter that we evaluated. Admission rate was analysed noting that admitted patients were primarily observed overnight.

## Statistical analysis

Data analysis was performed using SAS 9.3 basic descriptive statistics, such as means and standard deviations for continuous variables, and proportions and frequencies for categorical variables, were used to analyse patient characteristics. Continuous variables were grouped and converted to categorical variables for data analysis. Comparisons were made using two-sample t-test for continuous values, and Chi-square test/Fisher’s Exact Test for categorical variables. A p level of < 0.05 was used to determine statistical significance.

## Results

A total of 181 patients were identified within the specified time frame after application of the exclusion criteria. Of these, two additional patients were excluded since no data were recorded on their lactate levels. Patients were allocated into either the normal serum lactate group or elevated serum lactate group. Serum lactate levels are known to be affected by several of these. Seizure disorder was noted to be the foremost comorbid diagnosis in both groups. Alcoholism listed as a comorbidity is in reference to patients who have an alcoholic disorder but were not intoxicated at the time of evaluation. When the two groups were compared across all comorbidities, no statistical significance was noted.

The only variables statistically significant between the two groups were fluid use and Emergency Department length of stay (ED LOS) as illustrated in table-3 As would be expected, significantly more patients with elevated serum lactate received intravenous (IV) fluids than those in the normal serum lactate group (54.69% vs. 35.4%, p=0.01).

**: Table-3.** Statistical Significance between Variables.

| Variables | LA<2 mmol/L | LA>2 mmol/L | P-value |
| --- | --- | --- | --- |
| Shock Index (Range) | 1.5 (0.9-3) | 1.4 (0.9-2.6) | 0.0908 |
| Rate of Admission (%) | 9(7.8%) | 3 (4.7%) | 0.5419 |
| Rate of Readmission (72hr) | 8 (7.0%) | 7 (10.9%) | 0.3569 |
| ED LOS (minutes). | 255 (96-1169) | 195 (72-1335) | 0.0422* |
| BUN | 12 (3-48) | 11.5 (5-40) | 0.7982 |
| Cr | 0.8 (0.4-3.9) | 0.9 (0.4-1.8) | 0.5483 |
| Fluids (Liters) | 1 (0.1-2.3) | 1 (0.1-2.0) | 0.6838 |
| Rate of Fluids (Use) | 40 (35.4%) | 35 (54.69) | 0.0126* |
| Gender (M&F) | 50 (43.5%) | 29 (45.3%) | 0.8128 |
| Age (Range) | 40 (18-80) | 38 (18-82) | 0.5941 |
| Antibiotic Administration rate | 4 (3.9%) | 2 (3.2%) | 1 |
| # of Comorbidities | 1 (0-5) | 1 (0-5) | 0.3174 |
| Urine Drug Screen rate | 20 (19.8%) | 15 (25.9%) | 0.3747 |
| Urine Drug Screen-positive rate | 9 (50%) | 9 (64.29%) | 0.419 |
| Anti-epileptic meds use rate | 80 (72.1%) | 36 (58.1) | 0.0602 |

Cr: creatinine
ED LOS: emergency department length of stay
BUN: blood urea nitrogen

Interestingly, the median LOS was 60 minutes less in the elevated serum lactate group than in the normal lactate group (255 min vs. 195 min). The median calculation was used rather than the mean to avoid skewing the data secondary to extreme outliers. Anti-epileptic medication use was higher in the normal serum lactate group but did not reach statistical significance (72.1% vs 58.1%, p=0.06). No difference was found in the number of patients who had a drug screen performed; similarly, the number of patients with positive urine drug tests were not significantly different between those with normal serum lactate and those with elevated serum lactate (50% vs. 64.29%, p=0.4). Of note, two patients in the normal serum lactate group didn’t have a urine drug screen, hence the nine that were reported as positive in this group equated to a 50% positive rate. Positive results were reported for tetrahydrocannabinol (THC), amphetamines, opioids, benzodiazepines, ecstasy and cocaine. Renal function (BUN and creatinine (Cr)), shock index (heart rate/systolic blood pressure), rate of admission and rate of re-admission at 72 hours were all found to be similar between the two groups.

## Discussion

Several studies have demonstrated that elevated serum lactate is associated with higher morbidity and mortality in the setting of sepsis, shock, or trauma ^15, 18, 19^. In our study population, there was no mortality; and after accounting for comorbidities, no statistically significant difference between patients with normal serum lactate and those with elevated serum lactate as illustrated in figure-1. This wasn’t unexpected in that elevated lactic acid levels associated with seizure did not correlate with morbidity and mortality given the aforementioned pathophysiology of the transient nature of lactate production during seizure. This study reaffirms the understanding that lactic acidosis related to seizure self-resolves secondary to source control upon cessation of the event. The number of patients studied was significantly diminished after applying the exclusion criteria, which may have affected the power of the study and could explain why certain differences were not found between groups. Another limitation was that the length of the seizure and time of serum lactate collection in the ED could not be determined retrospectively. This may have had an effect on lab values as the half-life of lactate after grand mal seizure is approximately 1 hour ^11^.

It has been shown that shorter waiting times in the ED are associated with decreased mortality and increased patient satisfaction ^20-22^. Excessive patient wait times and disposition delays contribute to poor patient flow in the ED ^23^. Our study found a significantly higher ED LOS by 60 minutes, in patients with normal serum lactate. This may have been because clinicians were searching for alternative diagnoses as normal lactate levels are generally not seen in the setting of the acute tissue hypoxia and hypo-perfusion that seizures create. The current study found that a higher number of patients in the elevated serum lactate group received IV fluids, which was expected. These results outline possible areas of improvement with regard to mortality, patient satisfaction, and cost of care.

## Limitations

The obstacles we encountered during the research are sample size, the patients that we reviewed their records might not be a true representation of the entire state or country health wise as not all the ED in the state of West Virginia participated in the research. Furthermore, several other pre-existing health problems in out participants may play the role of confounding in evaluating the levels of lactate during acute seizure.

## Conclusion

Although there were no significant differences found with regard to morbidity and mortality between patients with normal or elevated serum lactate, we take this as further evidence to support the understanding that lactic acidosis secondary to seizure is a self-limited phenomenon, and that serum lactate level can be very useful in categorizing generalized tonic-clonic seizure from syncope and other forms of psychogenic non-epileptic seizures. If a patient presents to the ED with a seizure of unknown origin associated with increased lactate level, there should be a high suspicion for a generalized tonic-clonic seizure. Patients with normal serum lactate and with long LOS there is rapid normalization of serum lactate after a generalized seizure and so its value as a sole diagnostic tool for is limited. More so, serum lactate has time-dependent sensitivity and specificity as a diagnostic tool, its use could be compared with creatinine kinase and prolactin in a future prospective study to garner useful information that could be implemented for process improvement and cost containment strategies in the ED, and as a baseline for future projects

## Data Availability

All data produced in the present work are contained in the manuscript.

---

**Graph-1:**
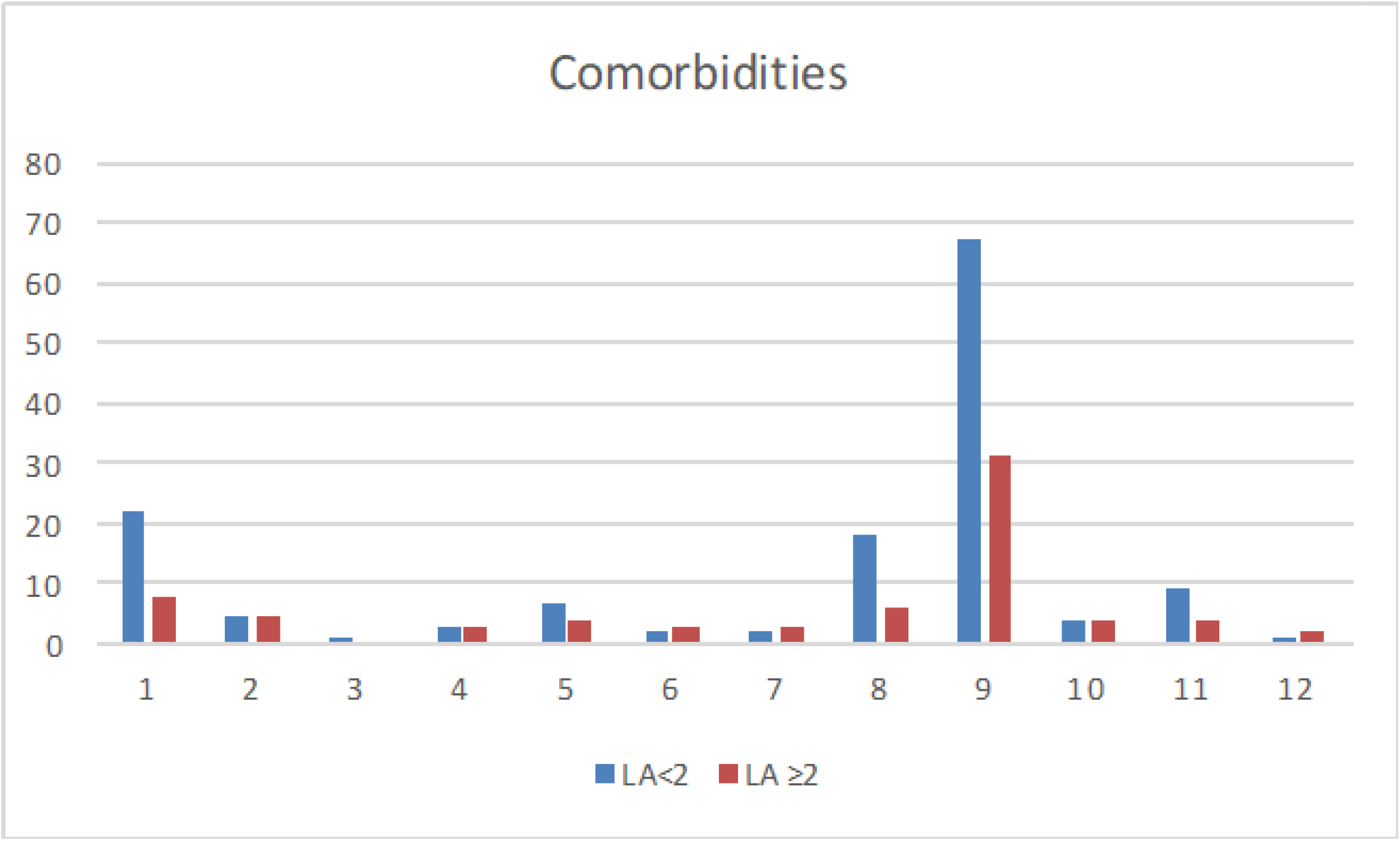
Comorbidities in LA groups. 1-HTN; 2-DM; 3-CHF; 4-COPD; 5-Active Malignancy; 6-CAD; 7-DVT; 8-Obesity; 9-Seizure Disorder; 10-Drug Addiction; 11-Alcoholism; 12-Other. **HTN:** Hypertension **DM:** Diabetes Mellitus **CHF:** Congestive Heart Failure **COPD:** Chronic Obstructive Pulmonary Disorder **CAD:** Coronary Artery Disease LA: lactic acid level in mmol/L

## Notes

### Competing Interest Statement

The authors have declared no competing interest.

### Funding Statement

This Study did not receive any funding.

### Author Declarations

Ethics committee/IRB of Charleston Area Medical center gave approval (IRB: 18-444) for this study.

